# Neuronal alpha-Synuclein Disease stage progression over five years

**DOI:** 10.1101/2024.12.17.24319172

**Authors:** Tanya Simuni, Caroline Gochanour, Anuprita R Nair, Michael C Brumm, Christopher Coffey, Kathleen L Poston, Lana M Chahine, Daniel Weintraub, Caroline M Tanner, Paulina Gonzalez-Latapi, Catherine M Kopil, Yuge Xiao, Sohini Chowdhury, Tien Dam, Gennaro Pagano, Diane Stephenson, Andrew Siderowf, Billy Dunn, Kenneth Marek, the Parkinson’s Progression Markers Initiative

## Abstract

**Background:** Neuronal alpha-Synuclein Disease (NSD) is defined by presence of an in vivo biomarker of neuronal alpha-synuclein (n-asyn) pathology, independent of presence of clinical syndrome. The NSD integrated staging system (NSD-ISS) describes progression across the disease continuum as stages 0 to 6. The objective of this analysis was to assess 5-year longitudinal change in the NSD-ISS.

**Methods:** Analysis included a subset of participants from the Parkinson’s Progression Markers Initiative (PPMI) enrolled before 2020 as Parkinson’s disease, prodromal, or healthy controls who met NSD criteria. Staging was defined based on biomarkers of n-asyn and dopaminergic dysfunction in early stages, clinical features (cognition, other non-motor features, and parkinsonism), and increasing degree of functional impairment in stages 3-6. Stages were examined annually for 5 years, along with the determinants of progression and effects of dopaminergic medication.

**Findings:** 576 participants were n-asyn positive and included in the analysis. Of these, 494 were enrolled as Parkinson’s disease, 74 as prodromal and 8 as healthy controls. At baseline 56% of participants were in stage 3, 24% stage 2B, 13% stage 4, 4% stage 2A, and <1% in the other stages. At year 5, the percent of individuals in stage 2B, 3, and 4 were 11%, 50%, and 34% respectively, indicating progression through NSD stages. Median (95% CI) time to next stage increase from baseline was 1·19 (1·07-2·01) for stage 2B, 4·98 (4·07-5·35) for stage 3, and 9·77 (7·04-NA) for stage 4. Progression was driven by functional impairment in predominantly motor domain (95%) for stage 2B to 3, increasing degree of non-motor dysfunction for stage 3 to 4 (46%), and combination of motor, non-motor and cognitive domains for stage 4 to 5. Initiation of dopaminergic medications lead to stage regression in 8% participants in stage 3 and 41% in stage 4.

**Interpretation:** Our analysis supports the validity of the NSD-ISS in defining stages of disease progression and support the utility of the NSD-ISS as a potential tool to identify clinical trial cohorts for drug development. Further validation in interventional studies is a crucial next step.

**Funding:** The Michael J. Fox Foundation for Parkinson’s Research (MJFF).

## INTRODUCTION

Neuronal alpha-Synuclein Disease (NSD) is defined by presence of an in vivo biomarker of neuronal alpha-synuclein (n-asyn) pathology.^1^ NSD encompasses Parkinson’s disease (PD), dementia with Lewy bodies (DLB), and any other n-asyn driven clinical syndrome. We have further proposed the Neuronal alpha-Synuclein Integrated Staging system (NSD-ISS) which integrates the biological substrates of the disease, n-asyn (S) and dopaminergic dysfunction (D), with cognitive, other non-motor, or motor manifestations and increasing degree of functional impairment to define stages along the NSD continuum. The intent of the NSD-ISS is to provide an integrated biological and clinical framework to expand our understanding of disease and advance biologically targeted therapeutic development.

The NSD-ISS proposed seven distinct stages: Stage 0 (presence of fully penetrant pathogenic variants in *SNCA* gene); Stage 1 [presence of n-asyn alone (Stage 1A) or in combination with dopaminergic dysfunction ( Stage 1B), asymptomatic]; Stage 2 [presence of n-asyn alone (Stage 2A) or in combination with dopaminergic dysfunction (Stage 2B), and subtle clinical signs/symptoms without functional impairment]; and Stages 3-6 [presence of both n-asyn and dopaminergic dysfunction, and clinical signs/symptoms with progressively increasing severity of functional impairment].^1^ We have subsequently developed biologic and clinical criteria and thresholds, utilizing currently available clinical and functional rating scales, to operationalize the NSD and NSD-ISS framework and have applied these definitions to assess cross-sectional baseline staging utilizing available baseline data in three well characterized studies.^2^ These results demonstrated that staging by NSD-ISS separated these early disease cohorts into NSD stages 2, 3 and 4 highlighting the significant heterogeneity in samples defined based on established clinical diagnostic criteria or features. The data further demonstrated that stage at baseline was a strong predictor of progression to a clinically meaningful milestones.^2^ However, the longitudinal stability of the NSD-ISS and the time for progression from one stage to the next have not been explored yet. These variables are key requirement for a disease staging system. The objectives of this analysis were to one, assess longitudinal change in NSD-ISS; two, domains of functional impairment (motor, non-motor, cognition) that lead to stage change; three, impact of PD medications on stability of the stage.

## METHODS

### Participants and Study Design

Data were from the Parkinson’s Progression Markers Initiative (PPMI) study. Study aims and methodology have been published elsewhere.^3–5^ Briefly, PPMI (NCT01141023) study is a multinational, prospective longitudinal observational study launched in 2010.^3^ Initial enrollment consisted of participants with early PD and healthy controls (HC). Subsequently, additional cohorts were added including participants with PD and non-manifesting carriers of genetic variants associated with PD, and participants with prodromal features (RBD or hyposmia). Individuals with PD were enrolled if they fulfilled PD diagnostic criteria, were within 2 years of diagnosis, Hoehn and Yahr (H&Y) stage 1–2, not on PD medications at the time of enrollment, and had an abnormal dopamine transporter (DAT) imaging scan with single-photon-emission computed tomography (SPECT). Inclusion criteria for the genetic PD cohort were the same, except for PD medications and diagnosis within 7 years were allowed. Prodromal participants had prodromal features associated with risk of PD, including severe hyposmia as measured by the University of Pennsylvania Smell Identification Test (UPSIT) based on internal population norms^6^ or REM sleep behavior disorder (RBD) confirmed by polysomnogram. Healthy controls were similar age and sex individuals without known neurological signs or symptoms and normal DAT imaging. For this analysis, only participants enrolled before 2020 (to allow the opportunity for at least 5 years of follow-up) who were n-asyn positive were included.

All PPMI participants undergo extensive clinical phenotypic and biological characterization annually. Participants undergo a series of clinical assessments described previously.^3–5^ Relevant assessments included the Montreal Cognitive assessment (MoCA)^7^ and the Movement Disorders Society Unified Parkinson’s Disease Rating Scale (MDS-UPDRS) parts I (non-motor aspects or experiences of daily living), II (motor aspects or experiences of daily living), and III (motor examination; recorded in the off**-**state at baseline for treated participants).^8^ Cerebrospinal fluid (CSF) samples are collected annually. Dopamine transporter (DAT) SPECT imaging is acquired longitudinally as per protocol.^9^

All study and recruitment materials were approved by institutional review boards or ethics committee at each site. Written informed consent was obtained from all participants before undergoing any study evaluations. The study was performed in accordance with the principles outlined in the Declaration of Helsinki and with Good Clinical Practice guidelines.

### Eligibility and Staging

NSD stage was assigned to each annual visit for all PPMI Participants enrolled prior to 2020 who were NSD positive and had sufficient data to determine stage. Participants were excluded if they had no CSF n-asyn available or if they did not have at least one follow-up visit. If participants attended a follow-up but were missing necessary data to determine their stage, data from the previous annual visit was carried forward. As previously published, stages were assigned according to change in DAT binding (over stages 2A to 2B), presence of non-motor and motor symptoms (over stages 0-6), and level of functional impairment as determined by components of MDS-UPRDS parts 1 and 2 (stages 3-6)^2^ (Supplementary Table 1).

### Statistical Analysis

NSD stage at baseline was tabulated for all participants and separately by enrollment cohort (PD, prodromal, HC) and subgroups (PD sporadic vs genetic, prodromal features (RBD, hyposmic)). Descriptive statistics at baseline were presented overall and by stage at baseline, including frequency (percentage) for categorical measures and mean and standard deviation (SD) for continuous measures. In cases when the mean/SD < 2 in the demographics tables, the median and interquartile range (IQR) were presented instead. Longitudinal analyses of change in stage was restricted to stage 2 and beyond due to the anticipated small sample size in stages 0-1B and excluded those who were in stage 2A and did not have at least one follow-up DAT SPECT scan. The frequency of NSD stage over time was tabulated at each annual visit through year 5 for all observed data. Stage over time was additionally presented for the subset of participants who completed a year 5 visit, using the last stage carried forward if participants did not have data for an interim visit. The first increase in stage from baseline, or “first progression,” was tabulated overall and by baseline stage, along with the mean (SD) and median (range) time to either the first progression in stage or the last visit. Participants in baseline stage groups with low counts were excluded and presented as supplementary material. The reason for progression from one stage to another, or so-called “track” was examined.

To further evaluate the relationship between baseline stage and stage progression, Kaplan-Meier plots were generated for time from enrollment to reaching the first stage increase, stratified by baseline stage. Time to reaching stage increase among those who were stage 2b at baseline was further stratified by enrollment cohort. Median time to stage increase and 95% confidence intervals are presented for each group. Participants who did not increase in stage or were lost to follow-up were right-censored. Time to first stage increase for these participants was calculated as the number of years from the enrollment date to the last follow-up visit date. Among those that progressed in stage from baseline, the tracks (cognitive, motor, and/or non-motor) that contributed to the stage increase were tabulated by stage at progression.

In order to assess the impact of symptomatic therapy on NSD stage over time, we plotted Kaplan-Meier survival curves of time from study enrolment to initiation of medication, stratified by baseline stage. Initiation of PD medication was defined as the first visit after a participant began actively taking PD medications (i.e. any medication contributing to the levodopa equivalency dose (LED) calculation).^10,11^ Participants who were on PD medication at baseline were excluded, as well as baseline stage groups with low numbers. Median time to initiating medication and 95% confidence intervals are presented for each group. Participants who did not initiate medication or were lost to follow-up were right-censored. Time to initiation of medication for censored participants was calculated as years from the enrollment date to the last follow-up visit date. Among those in stages 2B-4 at baseline who initiated PD medication, change in stage and key outcomes used to determine stage are presented for the last visit prior to initiating medication and the first visit on medication.

To evaluate study retention by baseline stage, Kaplan-Meier curves were generated for time to dropout, overall and stratified by stage. Time to study dropout was defined as time from enrollment to either withdrawal from the study or completion of the study per protocol at the 5 years visit. Participants who did not withdraw or complete the study were right censored at the last follow-up visit date. Median time to withdrawal or completion and 95% confidence intervals are presented overall and for each group.

Analyses were conducted using SAS v9·4 (SAS Institute Inc., Cary, NC; sas.com; RRID:SCR 008567). Sankey diagrams were created using “ggsankey” in R Statistical Software (v 4·3·2; R Core Team 2023).^12^

## RESULTS

576 participants were included in the analysis. See Figure 1 for the study flow chart and reasons for exclusion from the analysis. Table 1 provides baseline staging for the participants presented for the overall cohort and by enrollment cohorts and subgroup. For the overall cohort, 56% of individuals were in stage 3, defined by biomarkers of n-asyn and dopamine dysfunction and slight functional impairment, which is consistent with majority of the participants enrolled as early PD. There was substantial heterogeneity of staging at baseline in individuals recruited with the inclusion criteria of early sporadic PD (Table 1). While majority were in Stage 3, 24% were in stage 2B and 10% in stage 4. Participants recruited in the genetic PD subgroups, who were enrolled up to 7 years after diagnosis, had a higher percent in Stage 4. As expected, most individuals recruited into the RBD and Hyposmia subgroups of the prodromal cohort were stage 2 (presence of clinical signs but no functional impairment), although 13-17% were in stage 3. Only a minority of individuals recruited as non-manifesting carriers of relevant genetic variants (*GBA, LRRK2, SNCA*) fulfilled NSD criteria (Figure 1 and Table 1). Among those, the majority were in stages 2A/2B. Eight healthy controls (4% of those evaluable) fulfilled NSD criteria, stages 1-2.

**Figure 1.**
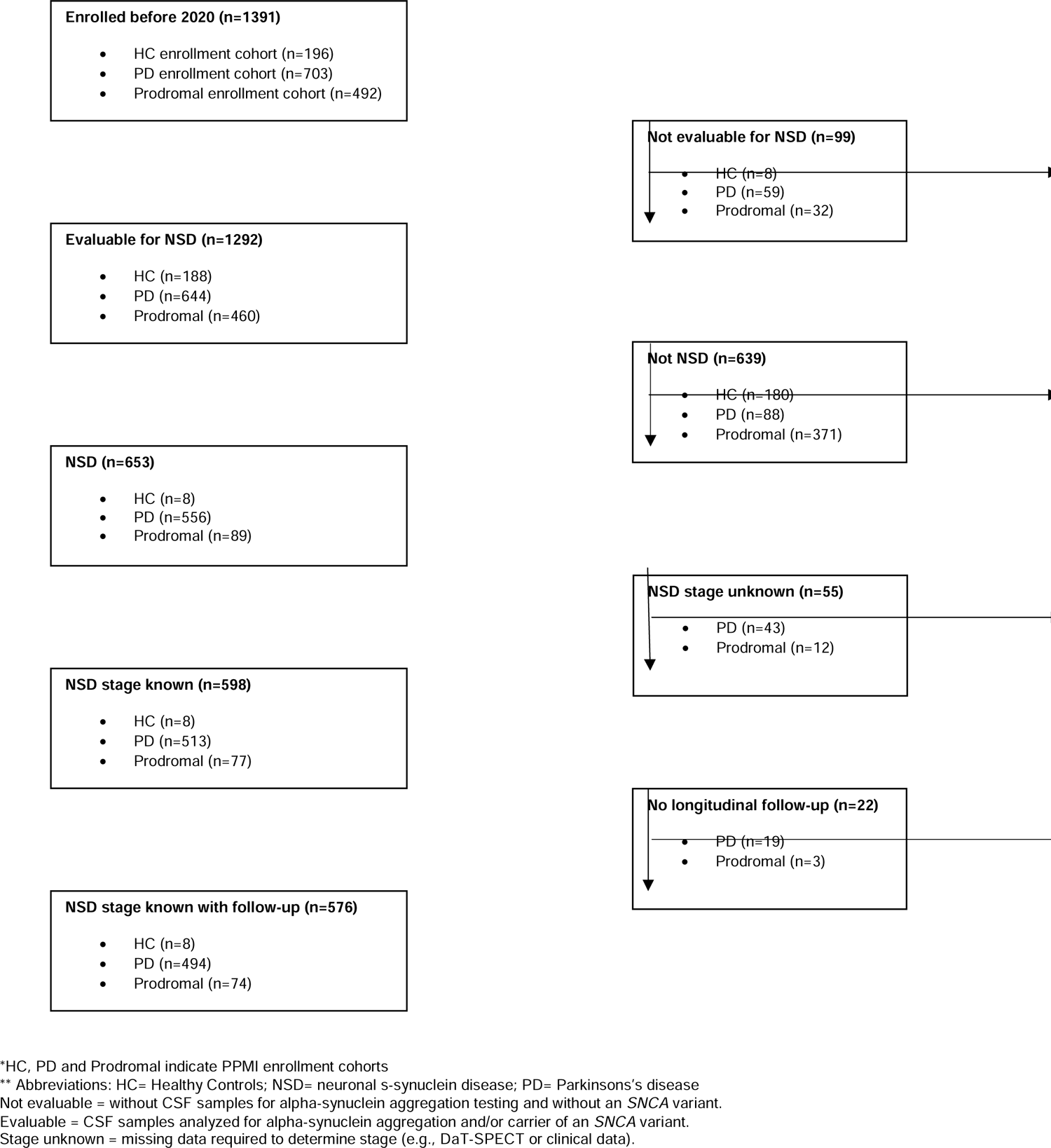
Flowchart

**Table 1.** Baseline staging of the PPMI NSD cohort.

| PPMI enrollment cohort |  |  |  |  |  |  |  |  |  |  |  |
| --- | --- | --- | --- | --- | --- | --- | --- | --- | --- | --- | --- |
|  |  | PD<br>(N=494) |  |  |  | Prodromal<br>(N=74) |  |  |  |  | HC<br>(N=8) |
| Stage | All<br>Participants<br>(N=576) | Sporadic<br>PD<br>(N=345) | LRRK2 PD*<br>(N=87) | GBA PD<br>(N=53) | SNCA PD<br>(N=9) | RBD<br>(N=30) | Hyposmia<br>(N=18) | LRRK2<br>NMC**<br>(N=15) | GBA NMC<br>(N=10) | SNCA<br>NMC<br>(N=1) | HC<br>(N=8) |
| Stage 0 | 1 (<1%) | — | — | — | 0 (NA) | — | — | — | — | 1 (NA) | — |
| Stage 1A | 10 (2%) | 0 | 0 | 0 | 0 (NA) | 0 | 0 | 4 (27%) | 3 (30%) | 0 (NA) | 3 (NA) |
| Stage 1B | 2 (<1%) | 0 | 0 | 0 | 0 (NA) | 0 | 0 | 2 (13%) | 0 | 0 (NA) | 0 (NA) |
| Stage 2A | 24 (4%) | 2 (1%) | 0 | 0 | 0 (NA) | 4 (13%) | 4 (22%) | 5 (33%) | 6 (60%) | 0 (NA) | 3 (NA) |
| Stage 2B | 137 (24%) | 83 (24%) | 12 (14%) | 7 (13%) | 1 (NA) | 18 (60%) | 11 (61%) | 3 (20%) | 0 | 0 (NA) | 2 (NA) |
| Stage 3 | 324 (56%) | 227 (66%) | 53 (61%) | 32 (60%) | 3 (NA) | 4 (13%) | 3 (17%) | 1 (7%) | 1 (10%) | 0 (NA) | 0 (NA) |
| Stage 4 | 73 (13%) | 33 (10%) | 20 (23%) | 13 (25%) | 4 (NA) | 3 (10%) | 0 | 0 | 0 | 0 (NA) | 0 (NA) |
| Stage 5 | 4 (1%) | 0 | 1 (1%) | 1 (2%) | 1 (NA) | 1 (3%) | 0 | 0 | 0 | 0 (NA) | 0 (NA) |
| Stage 6 | 1 (<1%) | 0 | 1 (1%) | 0 | 0 (NA) | 0 | 0 | 0 | 0 | 0 (NA) | 0 (NA) |

Data shown as n (%). Percentages displayed for subgroups with $\geq 10$ participants only. Abbreviations: HC = healthy controls; NMC = non-manifesting carriers; NSD = neuronal a-synuclein disease; NSD-ISS = neuronal a-synuclein disease integrated staging system; PD = Parkinson's disease; RBD = REM sleep behavior disorder; sPD = sporadic PD.
\* LRRK2 PD subgroup includes 3 participants who also carried a GBA mutation (2 in stage 3, 1 in stage 4).
\*\* LRRK2 NMC subgroup includes 2 participants who also carried a GBA mutation (1 in stage 1A, 1 in stage 2A).

Baseline demographic, clinical characteristics and biomarkers data of the participants with baseline NSD-ISS stages 2A-4 are presented in Table 2. Stages with less than 20 participants were excluded from the main table and instead presented as Supplementary Table 2. Notably, there was stage-dependent reduction of the DAT SPECT mean striatum specific binding ratio. 33% of participants had CSF aBeta 1-42 below the established cut off though there was no stage dependent difference. There was no stage dependent separation of the other reported biomarkers after accounting for multiple comparisons.

**Table 2.**
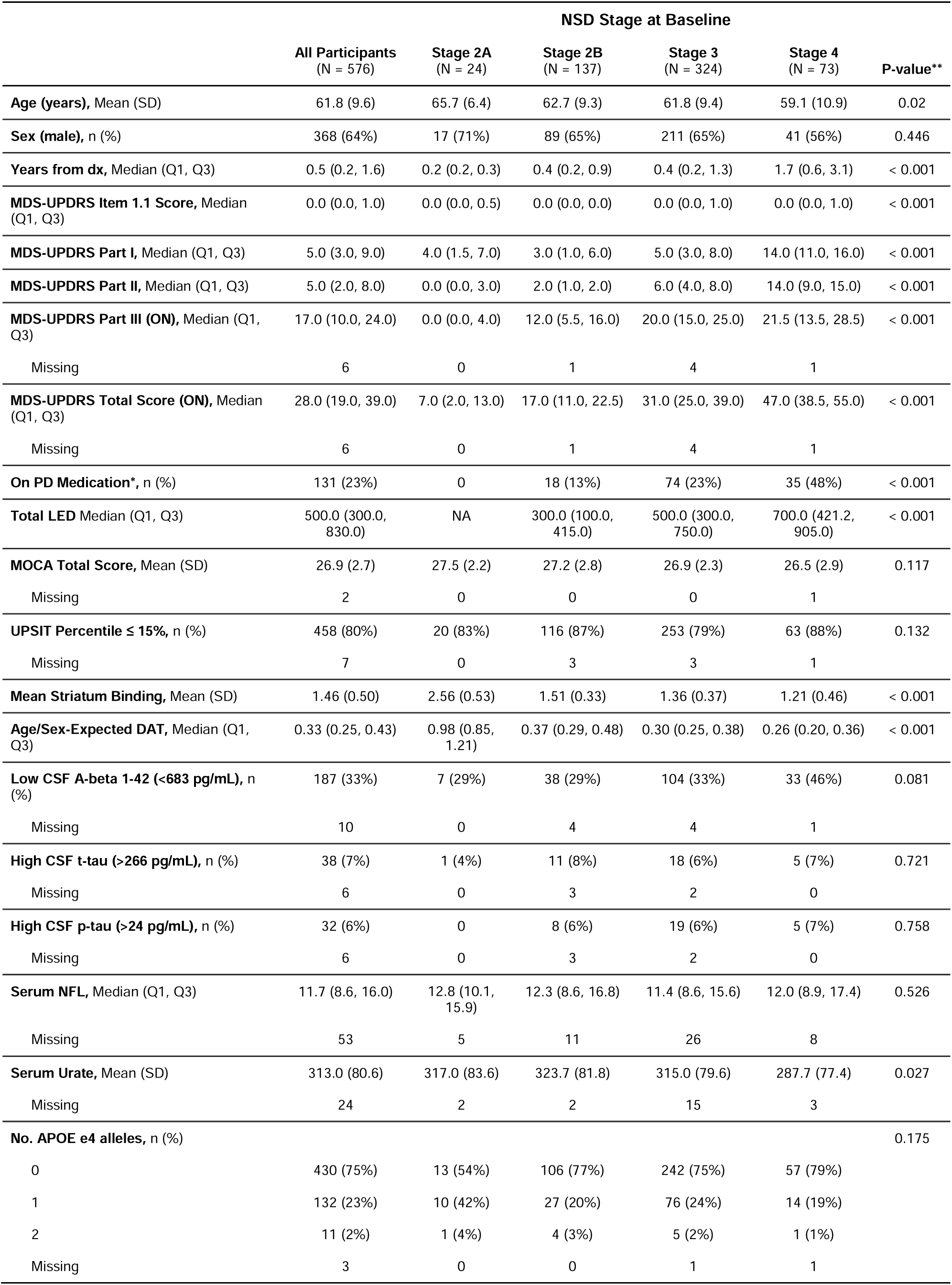
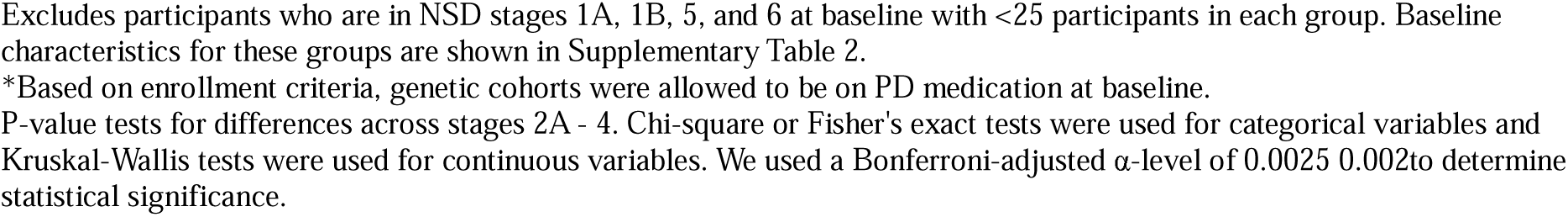
Clinical and biological baseline characteristics.

|  | NSD Stage at Baseline |  |  |  |  | P-value** |
| --- | --- | --- | --- | --- | --- | --- |
|  | All Participants<br>(N = 576) | Stage 2A<br>(N = 24) | Stage 2B<br>(N = 137) | Stage 3<br>(N = 324) | Stage 4<br>(N = 73) |  |
| **Age (years), Mean (SD)** | 61.8 (9.6) | 65.7 (6.4) | 62.7 (9.3) | 61.8 (9.4) | 59.1 (10.9) | 0.02 |
| **Sex (male), n (%)** | 368 (64%) | 17 (71%) | 89 (65%) | 211 (65%) | 41 (56%) | 0.446 |
| **Years from dx, Median (Q1, Q3)** | 0.5 (0.2, 1.6) | 0.2 (0.2, 0.3) | 0.4 (0.2, 0.9) | 0.4 (0.2, 1.3) | 1.7 (0.6, 3.1) | < 0.001 |
| **MDS-UPDRS Item 1.1 Score, Median (Q1, Q3)** | 0.0 (0.0, 1.0) | 0.0 (0.0, 0.5) | 0.0 (0.0, 0.0) | 0.0 (0.0, 1.0) | 0.0 (0.0, 1.0) | < 0.001 |
| **MDS-UPDRS Part I, Median (Q1, Q3)** | 5.0 (3.0, 9.0) | 4.0 (1.5, 7.0) | 3.0 (1.0, 6.0) | 5.0 (3.0, 8.0) | 14.0 (11.0, 16.0) | < 0.001 |
| **MDS-UPDRS Part II, Median (Q1, Q3)** | 5.0 (2.0, 8.0) | 0.0 (0.0, 3.0) | 2.0 (1.0, 2.0) | 6.0 (4.0, 8.0) | 14.0 (9.0, 15.0) | < 0.001 |
| **MDS-UPDRS Part III (ON), Median (Q1, Q3)** | 17.0 (10.0, 24.0) | 0.0 (0.0, 4.0) | 12.0 (5.5, 16.0) | 20.0 (15.0, 25.0) | 21.5 (13.5, 28.5) | < 0.001 |
| Missing | 6 | 0 | 1 | 4 | 1 |  |
| **MDS-UPDRS Total Score (ON), Median (Q1, Q3)** | 28.0 (19.0, 39.0) | 7.0 (2.0, 13.0) | 17.0 (11.0, 22.5) | 31.0 (25.0, 39.0) | 47.0 (38.5, 55.0) | < 0.001 |
| Missing | 6 | 0 | 1 | 4 | 1 |  |
| **On PD Medication\*, n (%)** | 131 (23%) | 0 | 18 (13%) | 74 (23%) | 35 (48%) | < 0.001 |
| **Total LED Median (Q1, Q3)** | 500.0 (300.0, 830.0) | NA | 300.0 (100.0, 415.0) | 500.0 (300.0, 750.0) | 700.0 (421.2, 905.0) | < 0.001 |
| **MOCA Total Score, Mean (SD)** | 26.9 (2.7) | 27.5 (2.2) | 27.2 (2.8) | 26.9 (2.3) | 26.5 (2.9) | 0.117 |
| Missing | 2 | 0 | 0 | 0 | 1 |  |
| **UPSIT Percentile ≤ 15%, n (%)** | 458 (80%) | 20 (83%) | 116 (87%) | 253 (79%) | 63 (88%) | 0.132 |
| Missing | 7 | 0 | 3 | 3 | 1 |  |
| **Mean Striatum Binding, Mean (SD)** | 1.46 (0.50) | 2.56 (0.53) | 1.51 (0.33) | 1.36 (0.37) | 1.21 (0.46) | < 0.001 |
| **Age/Sex-Expected DAT, Median (Q1, Q3)** | 0.33 (0.25, 0.43) | 0.98 (0.85, 1.21) | 0.37 (0.29, 0.48) | 0.30 (0.25, 0.38) | 0.26 (0.20, 0.36) | < 0.001 |
| **Low CSF A-beta 1-42 (<683 pg/mL), n (%)** | 187 (33%) | 7 (29%) | 38 (29%) | 104 (33%) | 33 (46%) | 0.081 |
| Missing | 10 | 0 | 4 | 4 | 1 |  |
| **High CSF t-tau (>266 pg/mL), n (%)** | 38 (7%) | 1 (4%) | 11 (8%) | 18 (6%) | 5 (7%) | 0.721 |
| Missing | 6 | 0 | 3 | 2 | 0 |  |
| **High CSF p-tau (>24 pg/mL), n (%)** | 32 (6%) | 0 | 8 (6%) | 19 (6%) | 5 (7%) | 0.758 |
| Missing | 6 | 0 | 3 | 2 | 0 |  |
| **Serum NFL, Median (Q1, Q3)** | 11.7 (8.6, 16.0) | 12.8 (10.1, 15.9) | 12.3 (8.6, 16.8) | 11.4 (8.6, 15.6) | 12.0 (8.9, 17.4) | 0.526 |
| Missing | 53 | 5 | 11 | 26 | 8 |  |
| **Serum Urate, Mean (SD)** | 313.0 (80.6) | 317.0 (83.6) | 323.7 (81.8) | 315.0 (79.6) | 287.7 (77.4) | 0.027 |
| Missing | 24 | 2 | 2 | 15 | 3 |  |
| **No. APOE e4 alleles, n (%)** |  |  |  |  |  | 0.175 |
| 0 | 430 (75%) | 13 (54%) | 106 (77%) | 242 (75%) | 57 (79%) |  |
| 1 | 132 (23%) | 10 (42%) | 27 (20%) | 76 (24%) | 14 (19%) |  |
| 2 | 11 (2%) | 1 (4%) | 4 (3%) | 5 (2%) | 1 (1%) |  |
| Missing | 3 | 0 | 0 | 1 | 1 |  |

\*Based on enrollment criteria, genetic cohorts were allowed to be on PD medication at baseline.
P-value tests for differences across stages 2A - 4. Chi-square or Fisher's exact tests were used for categorical variables and Kruskal-Wallis tests were used for continuous variables. We used a Bonferroni-adjusted $\alpha$ -level of 0.0025 to determine statistical significance.

Longitudinal staging of the cohort as a whole is presented in Figure 2. At the cohort level, there was reduction of the percent of individuals in stage 2B, relatively stable percent of individuals in stage 3 and increase in stage 4, reflecting expected progression across stages. A number of individuals did not progress across stages during the reported follow-up. For the individuals who progressed, majority advanced to the next stage, though 19/137 stage 2B progressors advanced to stage 4 (Table 3). Sensitivity analysis on 414 participants who had observed 5-year data had very similar results (Supplementary Figure 1). We further explored stage progression at the participant level including only 414 individuals who had 5 year observed data (Figure 3). Majority of participants with baseline stage 2B progressed to stage 3 with few “reversions back (Figure 3A). A higher proportion of participants with baseline stage 3 remained stable and those who progressed did not revert (Figure 3B). Majority of participants in baseline stage 4 remained in that stage, few advanced to stage 5/6, and a number reverted to stage 3 (Figure 3C).

**Figure 2.**
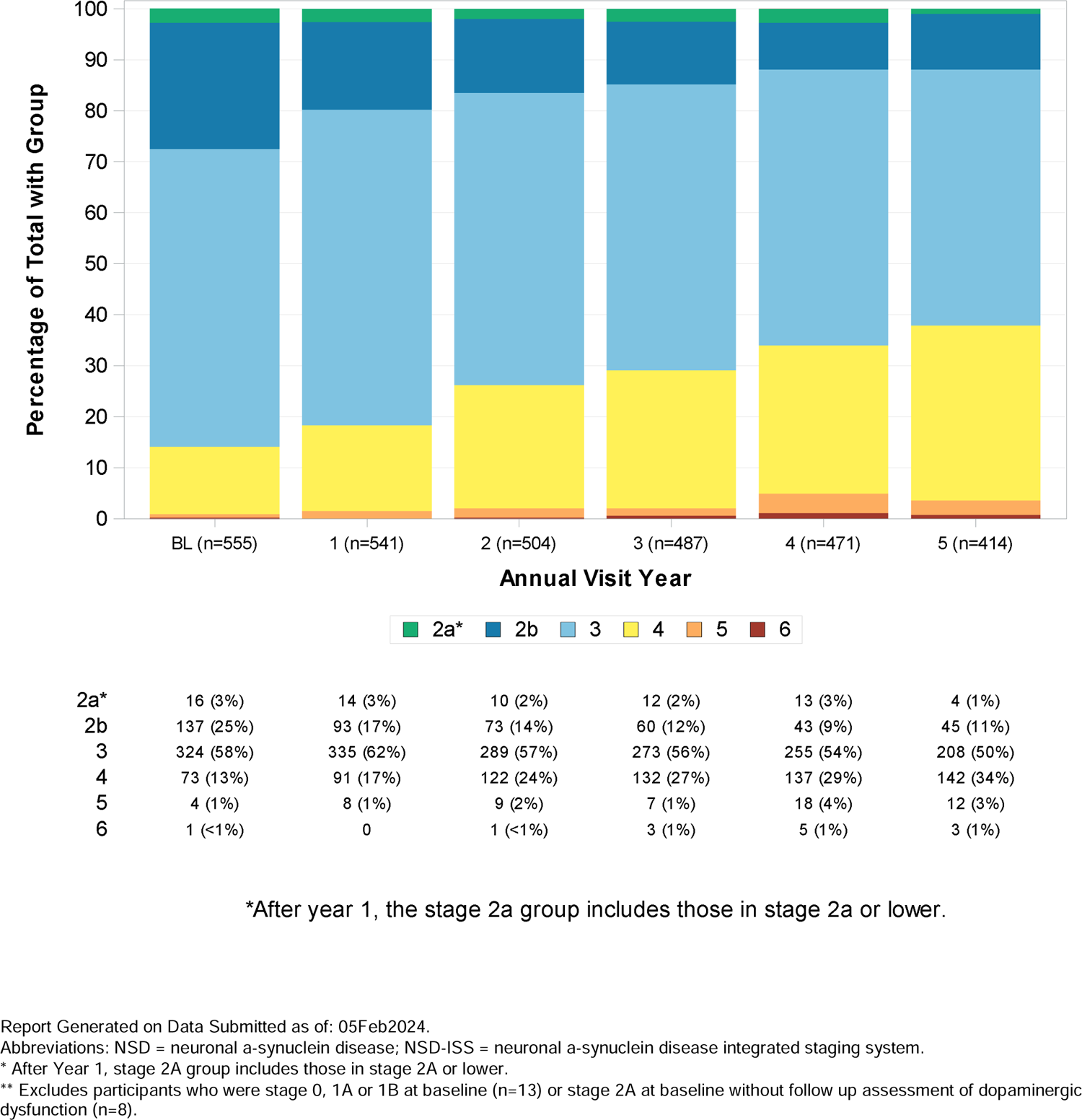
Longitudinal staging of the PPMI NSD cohort

**Figure 3.**
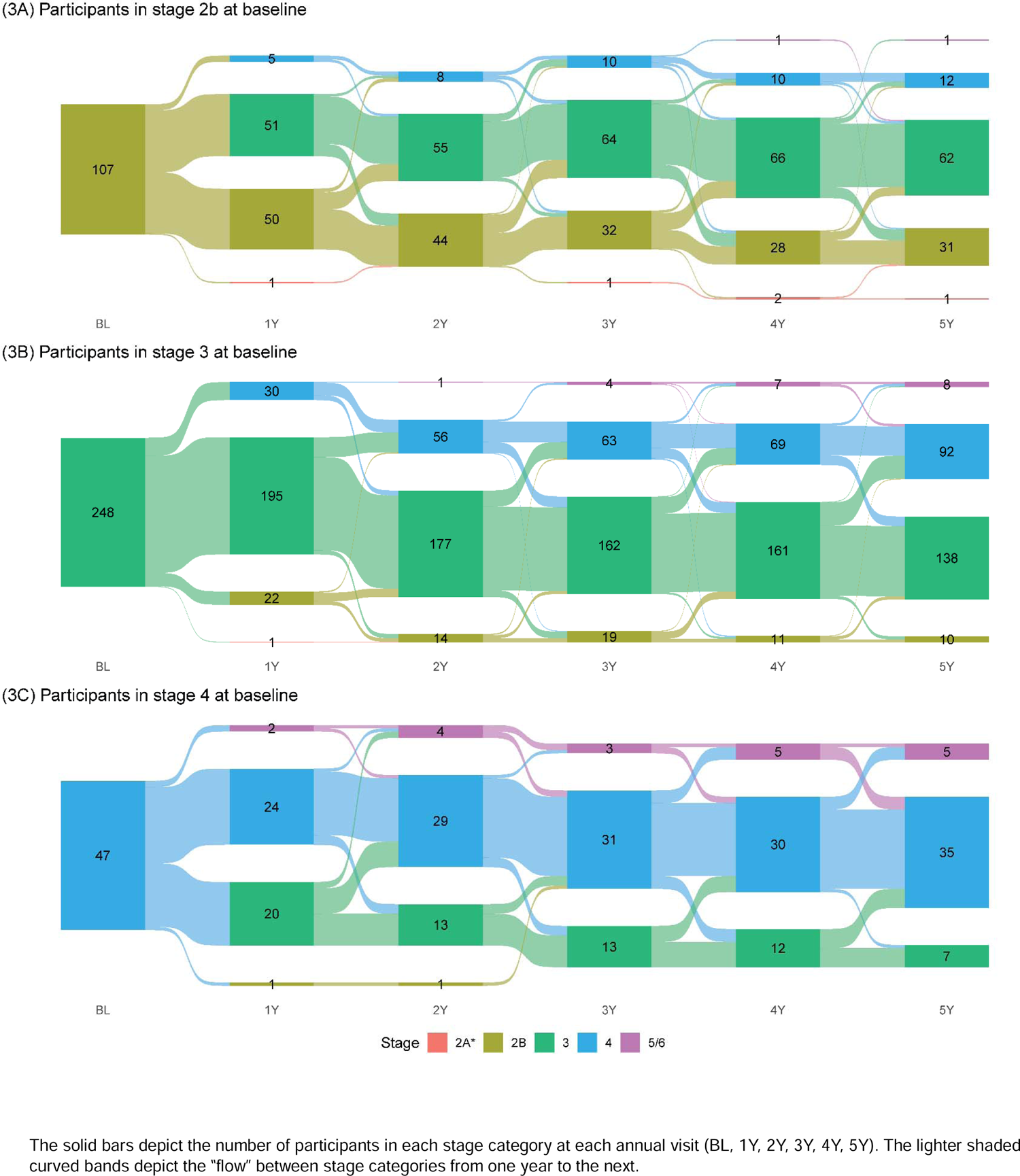
Participant flow

**Table 3.** First Stage Progression from Baseline.

|  | NSD Stage at Baseline |  |  |  |  |
| --- | --- | --- | --- | --- | --- |
|  | Overall<br>(N=550)* | Stage 2A<br>(N=16) | Stage 2B<br>(N=137) | Stage 3<br>(N=324) | Stage 4<br>(N=73) |
| **Years to first progression or last visit** |  |  |  |  |  |
| Mean (SD) | 4.2 (3.0) | 3.5 (2.5) | 2.4 (1.9) | 4.7 (3.1) | 5.3 (3.2) |
| Median (Min, Max) | 3.2 (0.8, 13.0) | 2.9 (1.0, 9.2) | 1.2 (0.8, 9.2) | 4.0 (0.8, 13.0) | 5.0 (1.0, 12.0) |
| **Stage at first progression, N** |  |  |  |  |  |
| No Progression | 150 | 7 | 10 | 87 | 46 |
| 2B | 6 | 6 | 0 | 0 | 0 |
| 3 | 110 | 2 | 108 | 0 | 0 |
| 4 | 255 | 1 | 19 | 235 | 0 |
| 5 | 28 | 0 | 0 | 2 | 26 |
| 6 | 1 | 0 | 0 | 0 | 1 |

\*Excludes participants in NSD stages 1A (n=5), 1B (n=2), 5 (n=4), and 6 (n=1) at baseline due to small numbers.

While these data do not allow us to determine the duration of each stage, data on time to first stage progression from baseline are presented in Table 3. Median (min, max) time from baseline to the first stage increase or last visit was 2·9 (1·0, 9·2) for stage 2A, 1·2 (0·8, 1·2) for stage 2B, 4·0 (0·8, 13·0) for stage 3 and 5 (1·0, 12·0) for stage 4. Figure 4A presents the time of progression from stage 2B to stage 3 or above. 50% of baseline stage 2B participants advanced to stage 3 or above by 1·19 (CI 1·07-2·01) years. The data are presented for all stage 2B participants and subdivided by the enrollment cohort (early PD versus prodromal). As expected, time to progression to stage 3 or above was longer in the latter (3·08 (1·07,4·16) versus 1·11 (1·06, 1·97)). Figure 4B presents the data on the time for stage increase for participants across stages 2B-4. Time to progression to the next stage increased with increasing stage.

**Figure 4A.**
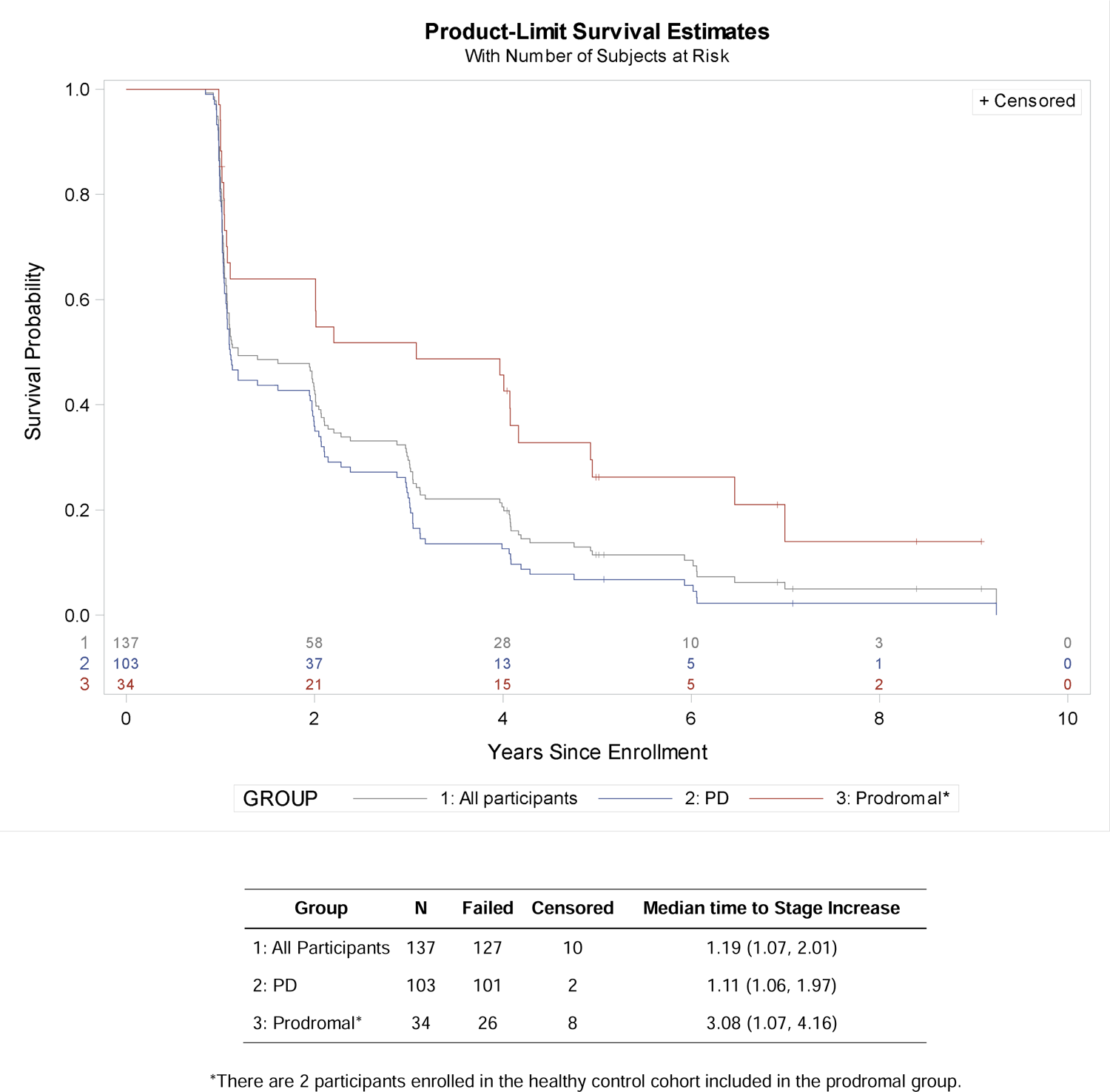
Time to reaching stage 3 and above among those who were stage 2B at baseline (PD vs Prodromal)

**Figure 4B.**
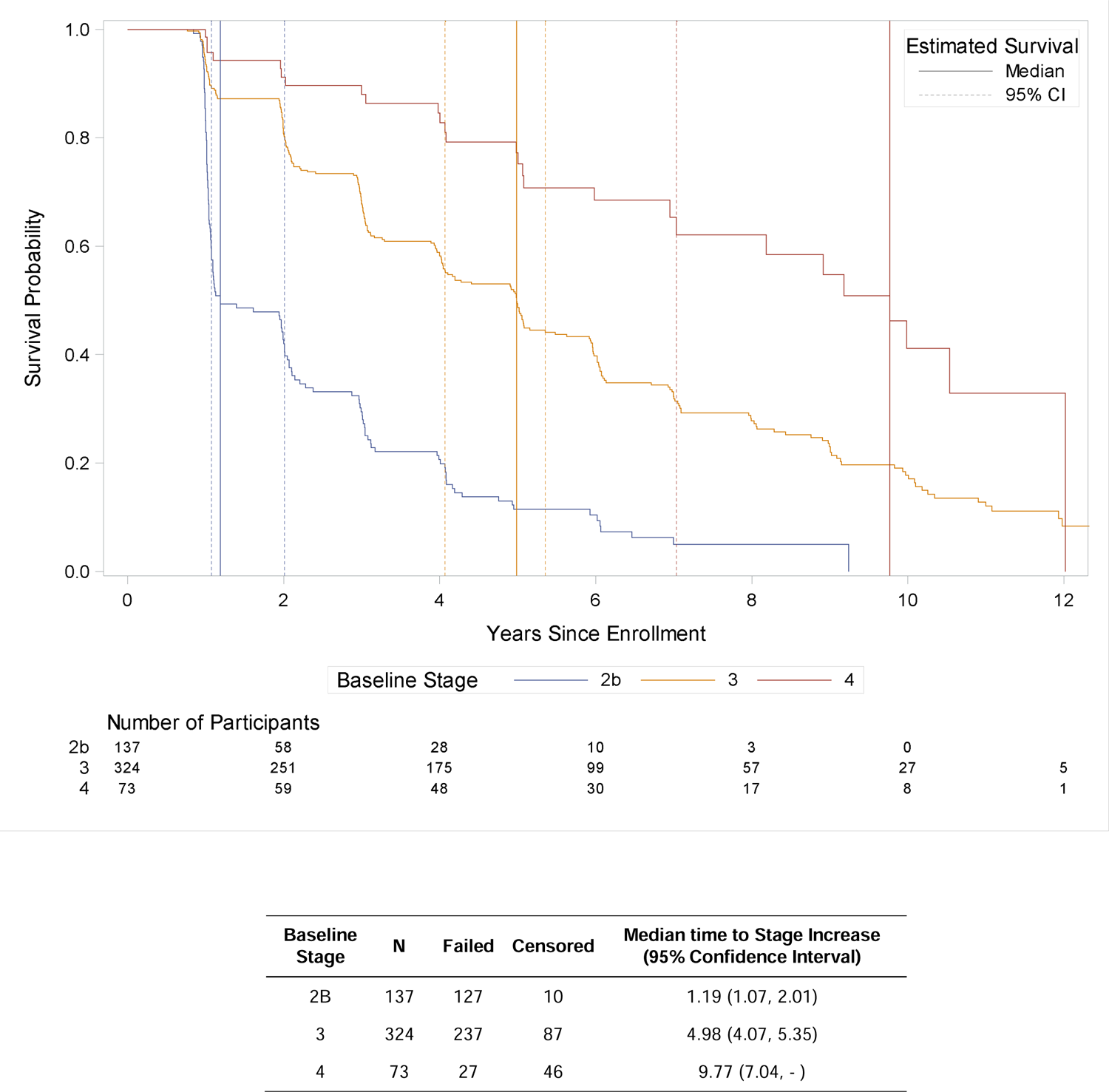
Time to reaching next stage increase by baseline stage

Figure 5 presents the data on the impact of baseline staging on time to initiation of PD medications. The analysis was restricted to individuals who were not taking such at baseline. The median (95% CI) time to initiation of PD medications was 2·05 (2·00-2·97) years in stage 2B, 1·09 (1·07-1·14) years in stage 3, and 1·07 (1·01-1·17) years in stage 4. We have further assessed the impact of initiation of PD medications on the stage stability and progression (Table 4). Importantly, there was no stage reversion (reduction) in individuals who progressed from stage 2B, and low number of reversions in baseline stage 3 (8%), while there was significant reversion (41%) in baseline stage 4. The majority of individuals in stage 2B progressed in stage despite initiation of PD medications, while for the majority of individuals in baseline stage 3, the stage remained stable (80%).

**Figure 5.**
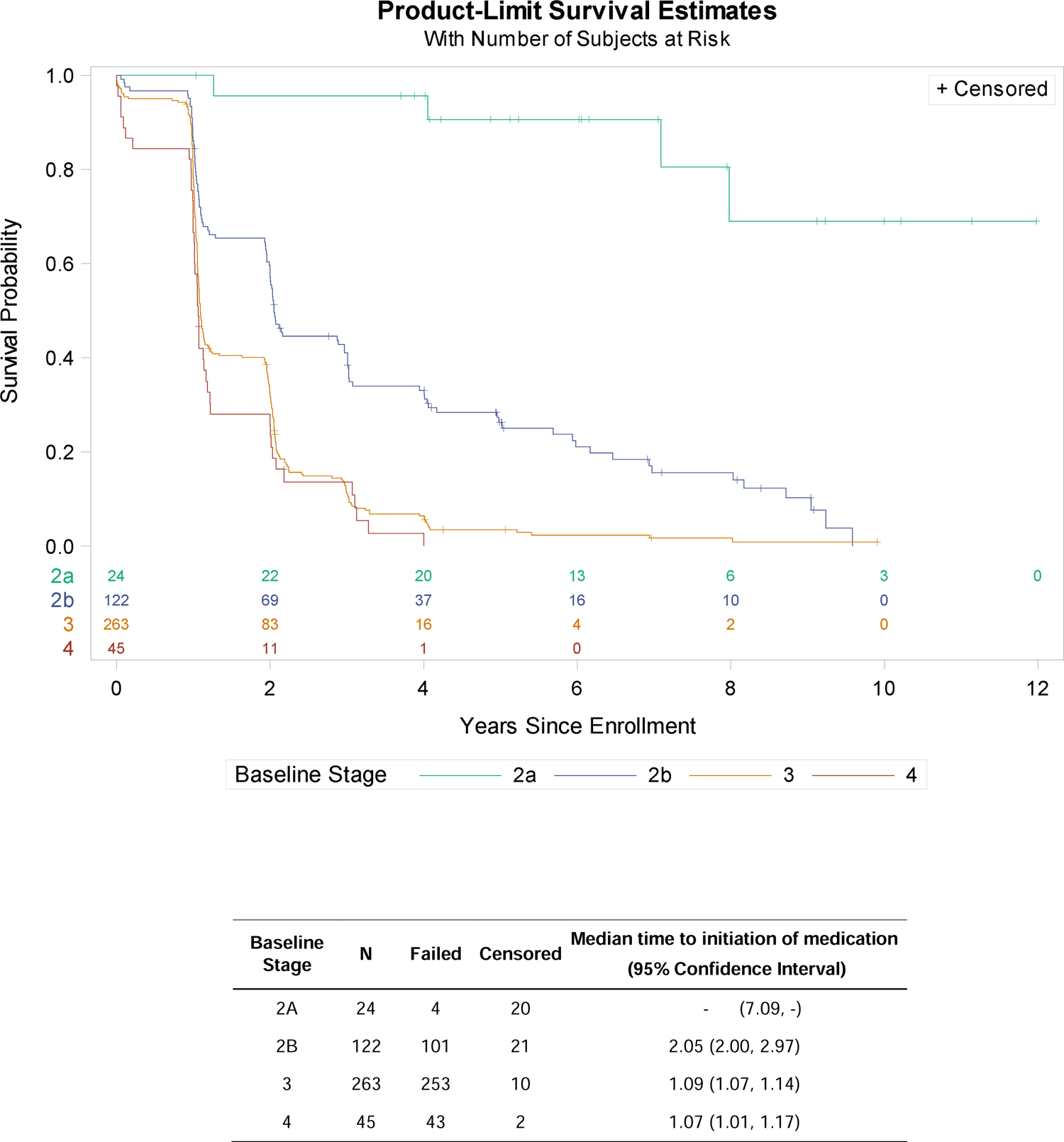
Time to initiation of PD medication by baseline stage (excluding those on medication at baseline)

**Figure 6.**
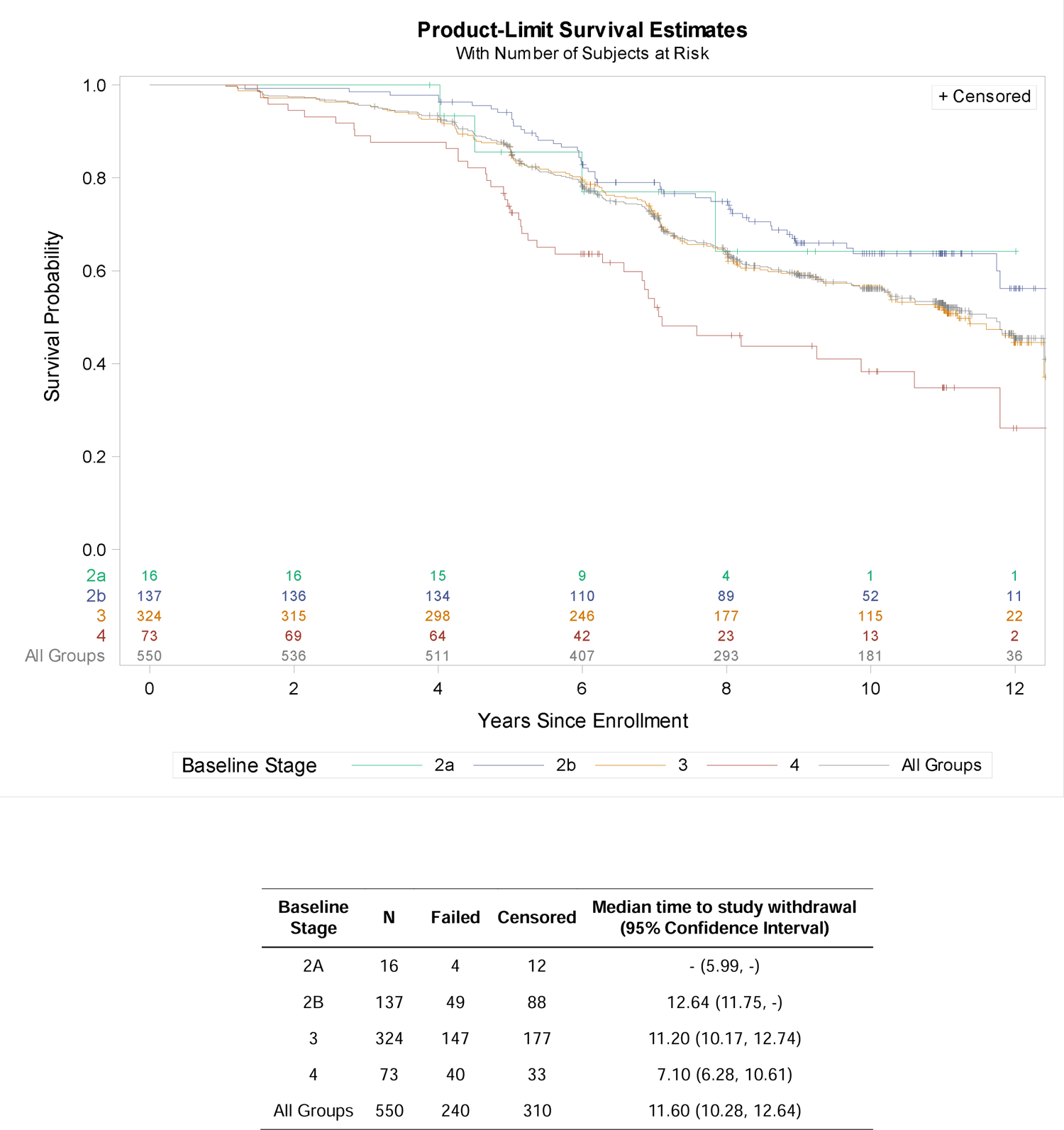
Time to study withdrawal

**Table 4.** Impact of PD medications on NSD stage and key outcomes at “last OFF medications visit” vs. “first ON medications visit” among participants who initiated ST *.

| Variable | Stage at last OFF visit |  |  |
| --- | --- | --- | --- |
|  | Stage 2B<br>(N=65) | Stage 3<br>(N=256) | Stage 4<br>(N=54) |
| Stage status at first ON visit, n (%) |  |  |  |
| Stable | 23 (35%) | 204 (80%) | 28 (52%) |
| Progressed | 42 (65%) | 31 (12%) | 4 (7%) |
| Reverted | NA | 21 (8%) | 22 (41%) |
| MDS-UPDRS Item 1.1 at last OFF visit | 0.1 (0.3) | 0.3 (0.5) | 0.7 (0.9) |
| MDS-UPDRS Item 1.1 at first ON visit | 0.2 (0.5) | 0.4 (0.6) | 0.7 (0.8) |
| MDS-UPDRS I subscore at last OFF visit | 2.8 (2.2) | 5.4 (3.0) | 12.2 (4.4) |
| MDS-UPDRS I subscore at first ON visit | 4.5 (2.9) | 6.4 (3.7) | 11.1 (5.4) |
| MDS-UPDRS II at last OFF visit | 1.6 (0.7) | 6.9 (3.0) | 13.9 (4.4) |
| MDS-UPDRS II at first ON visit | 4.7 (4.0) | 7.4 (4.1) | 12.6 (6.3) |
| MDS-UPDRS III at last OFF visit | 17.1 (9.6) | 24.4 (9.7) | 29.8 (12.1) |
| MDS-UPDRS III (ON) at first ON visit | 19.0 (11.1) | 22.4 (10.4) | 27.1 (12.5) |
| Missing | 5 | 19 | 4 |
| MoCA at last OFF visit | 27.3 (2.2) | 26.7 (2.6) | 26.6 (2.7) |
| MoCA at first ON visit | 26.8 (2.5) | 26.3 (3.1) | 25.8 (3.7) |
| Total LED at first ON visit | 265.3 (193.0) | 291.0 (211.3) | 352.8 (237.7) |

Data shown as mean (SD) or n (%). Unless otherwise specified, fewer than five participants in each group missed any one assessment. \* Note: the “first ON visit” was defined as the first visit at which a participant was actively taking symptomatic therapy (i.e., any medication contributing to LED calculation).
Data download: 05FEB2024

Considering that NSD-ISS provides anchors for stage progression across motor, cognitive and other non-motor domains, we assessed what “track” led to stage progression (Table 5). The majority of individuals progressing to stage 3, progressed on a purely motor “track” (91%), while individuals progressing to stage 4, advanced based on all three tracks though still with the higher proportion on the motor track. Progression to stage 5 was driven by cognitive domain in 29%, motor domain in 36%, non-motor domain in 11%, with the notable low number advancing based on combination of tracks. We ran similar analysis for the time to stage progression and tracks within 3 years from baseline (Supplementary Table 3).

**Table 5.** Tracks leading to stage progression.

| Track | Subgroup |  |  |
| --- | --- | --- | --- |
|  | Progressed to Stage<br>3<sup>a</sup><br>(N=110) | Progressed to Stage<br>4<sup>b</sup><br>(N=255) | Progressed to Stage<br>5<sup>c</sup><br>(N=28) |
| **Years to first progression, mean (SD)** | 2.1 (1.6) | 4.0 (2.7) | 5.0 (3.4) |
| Median (IQR) | 1.1 (1.0, 3.0) | 3.1 (2.0, 6.0) | 4.1 (2.0, 7.6) |
| **Met criteria for domain<sup>d</sup>** |  |  |  |
| Cognitive | 10 (9%) | 40 (16%) | 11 (39%) |
| Motor | 105 (95%) | 152 (60%) | 17 (61%) |
| Other Non-Motor | N/A | 117 (46%) | 8 (29%) |
| **Combination of domains** |  |  |  |
| Cognitive only | 5 (5%) | 27 (11%) | 8 (29%) |
| Motor only | 100 (91%) | 106 (42%) | 10 (36%) |
| Other Non-Motor only | N/A | 73 (29%) | 3 (11%) |
| Cognitive + Motor | 5 (5%) | 5 (2%) | 2 (7%) |
| Motor + Other Non-Motor | N/A | 36 (14%) | 4 (14%) |
| Cognitive + Other Non-Motor | N/A | 3 (1%) | 0 |
| Cognitive + Motor + Other Non-Motor | N/A | 5 (2%) | 1 (4%) |

<sup>a</sup> At baseline, 108 participants were in stage 2B and two were in stage 2A
<sup>b</sup> At baseline, 235 participants were in stage 3, 19 were in stage 2B, and one was in stage 2A.
<sup>c</sup> At baseline, 26 participants were in stage 4 and two were in stage 3.
<sup>d</sup> Indicates if criteria for corresponding domain were met irrespective of whether criteria for other domains were also met.

Lastly, we analyzed time to study withdrawal based on baseline stage. Analysis was restricted to individuals in stages 2A-4 at baseline (Figure 5). The risk of study withdrawal was higher in individuals in Stage 4 though, overall, more than 80% of the sample were retained in the study up to 5 years.

## DISCUSSION

We present the data on longitudinal change of NSD-ISS across the continuum of individuals with early PD, prodromal and healthy controls, recruited in PPMI study who fulfilled NSD criteria at baseline. We tracked stage changes in the PPMI cohort over 5 years of data analysis. Most participants either remained at their stage at baseline or progressed to the next higher stage during their observation period. Most individuals did not skip a stage or revert to a lower stage. Our focus was on Stages 2B, 3, and 4 where the majority of these participants were at baseline. Notably, there was progressive reduction of the number of individuals in Stage 2B and increase in stage 4, while the numbers in stage 3 remained relatively stable. This can be explained by expected progression of stage 2B to 3 and stage 3 to 4, supporting face validity of staging. Sensitivity analysis in individuals who had observed 5-year data demonstrated the same pattern.

Consistent with the previous report,^2^ we demonstrated significant baseline stage heterogeneity in individuals recruited into the study under the current definition of early PD. Such heterogeneity can have a significant impact on the read outs of disease modifying interventional studies as it adds to the variance of the cohort.

We also present the data on baseline clinical and biological characteristics of participants in stages 0-6, demonstrating stage dependent separation of a number of clinical variables that are not part of the staging anchors. Importantly, we demonstrate stage dependent separation of DAT imaging characteristics. These observations support face validity of the staging construct though additional investigation of the biological markers in stage dependent progression is necessary and underway.

Since the majority of these data are from the original PPMI cohort enrolled as early PD (not by stage), it is unclear how long these individuals have been in their stage at baseline. Therefore, while these data are useful in describing heterogeneity of the baseline staging, they do not allow us to accurately determine the duration of these participants within a stage. However, we have provided data for this cohort for the time to first stage progression from baseline. These data suggest that for this early PD and prodromal cohort, the change in early stages (stage 2B and 3) are within the timeframe acceptable for the design of interventional studies. As an example, 50% of individuals progressed from stage 2B to 3 by median 1·19 (1·07; 2·01) years from recruitment, and 75% by median 3·08 (2·87, 4·08) years from recruitment. Such a time frame can make stage progression a viable outcome for disease modification studies. These results have to be taken with caution as individuals recruited into this cohort might represent “the tail end” of Stage 2B. Further investigation in a cohort followed from the start of stage 2B is underway in PPMI.

The initiation of PD medications in clinical trials targeting early PD has resulted in a key confound for interpretation of longitudinal data such as the MDS-UPDRS. Most studies target individuals who do not take PD medications and censor the data once such were initiated. An alternative approach piloted recently is time to event analysis that has been demonstrated to be less susceptible to initiation of PD medications.^13^ While we envision that NSD-ISS provides the framework to start interventional studies in earlier stages when PD medications are not relevant, our data provide a number of important observations supporting relative “resistance” of NSD-ISS, specifically in early stages (stage 2B and 3), to the effect of PD medications. Our data also provide another rationale to exclude individuals in Stage 4 from disease modifying trials due to faster time to initiation of PD medications, higher dropout rate and higher percent of stage reversal after initiation of PD medications. Proposed NSD-ISS anchors allow to easily stage individuals as part of the inclusion criteria, provided biomarker data are available.

Lastly, we analyzed impact of baseline stage on time to study withdrawal. Not surprisingly, individuals in stage 4 had shorter time to withdrawal reflecting increasing functional impairment. However, important for studies design, most of the withdrawals occurred post 5 years of observation indicating it as a less relevant issue for the studies with traditional duration of one-three years.

In summary, the strength of our data is the comprehensive ascertainment of stage progression in a large cohort of biologically defined and deeply phenotypically characterized individuals. The study has a number of limitations that serve to set the objectives for future research. Most importantly, the majority of the cohort that had 5+ years of longitudinal data were recruited as early PD and as expected, fall into predominantly stage 3. While 24% of the cohort were in stage 2B, most of them still had PD clinical phenotype (but no functional impairment). Less than 10% of the cohort were in stage 2A or below. As therapeutic development is moving into disease prevention, the data on timelines and predictors of stage 1 and 2A progression are essentially necessary. Identification of stage 1 cohorts (strictly biomarkers defined) will require population screening and will be predicated on availability of feasible and scalable biomarkers; The field is actively working on such, and we hope that such cohorts will be launched in the next 2-5 years. Identification of stage 2A individuals is feasible with currently available tools, and PPMI is aiming to recruit about 1000 such participants in the next 5 years. Another limitation is that currently reported PPMI cohort does not include individuals with DLB or prodromal DLB and has a limited number of individuals with stage 2B RBD phenotype. Since 2020, we have recruited a significant number of RBD stage 2A and 2B individuals and will be reporting longitudinal data as the cohort matures.

In conclusion, our data supports face validity of the NSD-ISS staging, provides data on the timeline of stages progression, and addresses a number of questions that are important to facilitate implementation of NSD-ISS as a tool for therapeutic development. We hope that these data will encourage application of NSD-ISS in other cohorts, including cohorts with baseline non-motor phenotypes to inform the field. Validation in interventional studies will be essentially important.

## Data Availability

Data used in the preparation of this article were obtained on February 5, 2024 from the Parkinson’s Progression Markers Initiative (PPMI) database (www.ppmi-info.org/access-data-specimens/download-data), RRID:SCR_006431. For up-to-date information on the study, visit www.ppmi-info.org., Protocol information for The Parkinson’s Progression Markers Initiative (PPMI) Clinical - Establishing a Deeply Phenotyped PD Cohort AM 3.2. can be found on protocols.io or by following this link: https://dx.doi.org/10.17504/protocols.io.n92ldmw6ol5b/v2.

## ACKNOWLEDGEMENTS

We appreciate Ms. Tali Cohen, MA support for editorial review of the paper.

## Data Tier Information

This analysis was conducted by the PPMI Statistics Core and used actual dates of activity for participants, a restricted data element not available to public users of PPMI data.

## CODE AVAILABILITY STATEMENT

Statistical analysis codes used to perform the analyses in this article are shared on Zenodo [link].

## AUTHOR CONTRIBUTIONS

All authors contributed to conceptualization, investigations, methodology, and supervision. TS, CG, AN, MB, KP, LC, DW, PG-L, GP, and AS contributed to the review and editing of the final version of the manuscript. TS, CG, AN, MB, CC, LC, AS, BD, and KM contributed to data curation, formal analysis, and validation. TS, CG, AN, MB, CC, LC, DW, PG-L, TD, GP, AS, and KM contributed to the writing of the original draft. TS, CK, YX, and SC contributed to the project administration, acquired funding, and provided resources. All authors had direct access to and verified the underlying data.

## PPMI STUDY TEAMS/CORES/COLLABORATORS FOR PUBLICATIONS

### Executive Steering Committee

Kenneth Marek^1^; Caroline Tanner^9^; Tanya Simuni, MD^3^; Andrew Siderowf^12^; Douglas Galasko^27^;LJLana Chahine^39^; Christopher Coffey^4^; Kalpana Merchant^59^; Kathleen Poston^38^; Roseanne Dobkin^41^; Tatiana Foroud^15^; Brit Mollenhauer^8^; Dan Weintraub^12^; Ethan Brown^9^; Karl Kieburtz^23^; Mark Frasier^6^; Todd Sherer^6^; Sohini Chowdhury^6^; Roy Alcalay^35^ and Aleksandar Videnovic^45^

### Steering Committee

Duygu Tosun-Turgut^9^; Werner Poewe^7^; Susan Bressman^14^; Jan Hammer^15^; Raymond James^22^; Ekemini Riley^40^; John Seibyl^1^; Leslie Shaw^12^; David Standaert^18^; Sneha Mantri^60^; Nabila Dahodwala^12^; Michael Schwarzschild^45^; Connie Marras^43^; Hubert Fernandez^25^; Ira Shoulson^23^; Helen Rowbotham^2^; Paola Casalin^11^ and Claudia Trenkwalder^8^

### Michael J. Fox Foundation (Sponsor)

Todd Sherer; Sohini Chowdhury; Mark Frasier; Jamie Eberling; Katie Kopil; Alyssa O’Grady; Maggie McGuire Kuhl; Leslie Kirsch and Tawny Willson

### Study Cores, Committees and Related Studies

Project Management Core: Emily Flagg^1^

Site Management Core: Tanya Simuni^3^; Bridget McMahon^1^ Strategy and Technical Operations: Craig Stanley^1^; Kim Fabrizio^1^ Data Management Core: Dixie Ecklund^4^; Trevis Huff^4^

Screening Core: Tatiana Foroud^15^; Laura Heathers^15^; Christopher Hobbick^15^; Gena Antonopoulos^15^

Imaging Core: John Seibyl^1^; Kathleen Poston^38^

Statistics Core: Christopher Coffey^4^; Chelsea Caspell-Garcia^4^; Michael Brumm^4^

Bioinformatics Core: Arthur Toga^10^; Karen Crawford^10^

Biorepository Core: Tatiana Foroud^15^; Jan Hamer^15^

Biologics Review Committee: Brit Mollenhauer^8^; Doug Galasko^27^; Kalpana Merchant^59^

Genetics Core: Andrew Singleton^13^

Pathology Core: Tatiana Foroud^15^; Thomas Montine^38^

Found: Caroline Tanner^9^

PPMI Online: Carlie Tanner^9^; Ethan Brown^9^; Lana Chahine^39^; Roseann Dobkin^41^; Monica Korell^9^

### Site Investigators

Charles Adler^49^; Roy Alcalay^35^; Amy Amara^50^; Paolo Barone^30^; Bastiaan Bloem^58^ Susan Bressman^14^; Kathrin Brockmann^26^; Norbert Brüggemann^57^; Lana Chahine^39^; Kelvin Chou^42^; Nabila Dahodwala^12^; Alberto Espay^32^; Stewart Factor^16^; Hubert Fernandez^25^; Michelle Fullard^50^; Douglas Galasko^27^; Robert Hauser^19^; Penelope Hogarth^17^; Shu-Ching Hu^21^; Michele Hu^56^; Stuart Isaacson^31^; Christine Klein^57^; Rejko Krueger^2^; Mark Lew^47^; Zoltan Mari^54^; Connie Marras^43^; Maria Jose Martí^33^; Nikolaus McFarland^52^; Tiago Mestre^44^; Brit Mollenhauer^8^; Emile Moukheiber^28^; Alastair Noyce,^61^ Wolfgang Oertel^62^; Njideka Okubadejo^63^; Sarah O’Shea^37^; Rajesh Pahwa^46^; Nicola Pavese^55^; Werner Poewe^7^; Ron Postuma^3^; Giulietta Riboldi^51^; Lauren Ruffrage^18^; Javier Ruiz Martinez^34^; David Russell^1^; Marie H Saint-Hilaire^22^; Neil Santos^49^; Wesley Schlett^45^; Ruth Schneider^23^; Holly Shill^48^; David Shprecher^24^; Tanya Simuni^3^; David Standaert^18^; Leonidas Stefanis^36^; Yen Tai^29^; Caroline Tanner^9^; Arjun Tarakad^20^; Eduardo Tolosa^33^ and Aleksandar Videnovic^45^

### Coordinators

Susan Ainscough^30^; Courtney Blair^18^; Erica Botting^19^; Isabella Chung^54^; Kelly Clark^24^; Ioana Croitoru^34^; Kelly DeLano^32^; Iris Egner^7^; Fahrial Esha^51^; May Eshel^35^; Frank Ferrari^42^; Victoria Kate Foster^55^; Alicia Garrido^33^; Madita Grümmer^57^; Bethzaida Herrera^48^; Ella Hilt^26^; Chloe Huntzinger^50^; Raymond James^22^; Farah Kausar^9^; Christos Koros^36^; Yara Krasowski^58^; Dustin Le^17^; Ying Liu, MD^50^; Taina M. Marques^2^; Helen Mejia Santana^37^; Sherri Mosovsky^39^; Jennifer Mule^25^; Philip Ng^43^; Lauren O’Brien^46^; Abiola Ogunleye^29^; Oluwadamilola Ojo^63^; Obi Onyinanya^28^; Lisbeth Pennente^31^; Romina Perrotti^53^; Michael Pileggi^53^; Ashwini Ramachandran^12^; Deborah Raymond^14^; Jamil Razzaque^56^; Shawna Reddie^44^; Kori Ribb^28^; Kyle Rizer^52^; Janelle Rodriguez^27^; Stephanie Roman^1^; Clarissa Sanchez^20^; Cristina Simonet^29^; Anisha Singh^23^; Elisabeth Sittig^62^; Barbara Sommerfeld^16^; Angela Stovall^42^; Bobbie Stubbeman^32^; Alejandra Valenzuela^47^; Catherine Wandell^21^; Diana Willeke^8^; Karen Williams^3^ and Dilinuer Wubuli^43^

### Partners Scientific Advisory Board (Acknowledgement)

Funding: PPMI – a public-private partnership – is funded by the Michael J. Fox Foundation for Parkinson’s Research and funding partners, including 4D Pharma, Abbvie, AcureX, Allergan, Amathus Therapeutics, Aligning Science Across Parkinson’s, AskBio, Avid Radiopharmaceuticals, BIAL, BioArctic, Biogen, Biohaven, BioLegend, BlueRock Therapeutics, Bristol-Myers Squibb, Calico Labs, Capsida Biotherapeutics, Celgene, Cerevel Therapeutics, Coave Therapeutics, DaCapo Brainscience, Denali, Edmond J. Safra Foundation, Eli Lilly, Gain Therapeutics, GE HealthCare, Genentech, GSK, Golub Capital, Handl Therapeutics, Insitro, Jazz Pharmaceuticals, Johnson & Johnson Innovative Medicine, Lundbeck, Merck, Meso Scale Discovery, Mission Therapeutics, Neurocrine Biosciences, Neuron23, Neuropore, Pfizer, Piramal, Prevail Therapeutics, Roche, Sanofi, Servier, Sun Pharma Advanced Research Company, Takeda, Teva, UCB, Vanqua Bio, Verily, Voyager Therapeutics, the Weston Family Foundation and Yumanity Therapeutics.

1. Institute for Neurodegenerative Disorders, New Haven, CT
2. University of Luxembourg, Luxembourg
3. Northwestern University, Chicago, IL
4. University of Iowa, Iowa City, IA
5. VectivBio AG
6. The Michael J. Fox Foundation for Parkinson’s Research, New York, NY
7. Innsbruck Medical University, Innsbruck, Austria
8. Paracelsus-Elena Klinik, Kassel, Germany
9. University of California, San Francisco, CA
10. Laboratory of Neuroimaging (LONI), University of Southern California
11. BioRep, Milan, Italy
12. University of Pennsylvania, Philadelphia, PA
13. National Institute on Aging, NIH, Bethesda, MD
14. Mount Sinai Beth Israel, New York, NY
15. Indiana University, Indianapolis, IN
16. Emory University of Medicine, Atlanta, GA
17. Oregon Health and Science University, Portland, OR
18. University of Alabama at Birmingham, Birmingham, AL
19. University of South Florida, Tampa, FL
20. Baylor College of Medicine, Houston, TX
21. University of Washington, Seattle, WA
22. Boston University, Boston, MA
23. University of Rochester, Rochester, NY
24. Banner Research Institute, Sun City, AZ
25. Cleveland Clinic, Cleveland, OH
26. University of Tübingen, Tübingen, Germany
27. University of California, San Diego, CA
28. Johns Hopkins University, Baltimore, MD
29. Imperial College of London, London, UK
30. University of Salerno, Salerno, Italy
31. Parkinson’s Disease and Movement Disorders Center, Boca Raton, FL
32. University of Cincinnati, Cincinnati, OH
33. Hospital Clinic of Barcelona, Barcelona, Spain
34. Hospital Universitario Donostia, San Sebastian, Spain
35. Tel Aviv Sourasky Medical Center, Tel Aviv, Israel
36. National and Kapodistrian University of Athens, Athens, Greece
37. Columbia University Irving Medical Center, New York, NY
38. Stanford University, Stanford, CA
39. University of Pittsburgh, Pittsburgh, PA
40. Center for Strategy Philanthropy at Milken Institute, Washington D.C.
41. Rutgers University, Robert Wood Johnson Medical School, New Brunswick, New Jersey
42. University of Michigan, Ann Arbor, MI
43. Toronto Western Hospital, Toronto, Canada
44. The Ottawa Hospital, Ottawa, Canada
45. Massachusetts General Hospital, Boston, MA
46. University of Kansas Medical Center, Kansas City, KS
47. University of Southern California, Los Angeles, CA
48. Barrow Neurological Institute, Phoenix, AZ
49. Mayo Clinic Arizona, Scottsdale, AZ
50. University of Colorado, Aurora, CO
51. NYU Langone Medical Center, New York, NY
52. University of Florida, Gainesville, FL
53. Montreal Neurological Institute and Hospital/McGill, Montreal, QC, Canada
54. Cleveland Clinic-Las Vegas Lou Ruvo Center for Brain Health, Las Vegas, NV
55. Clinical Ageing Research Unit, Newcastle, UK
56. John Radcliffe Hospital Oxford and Oxford University, Oxford, UK
57. Universität Lübeck, Luebeck, Germany
58. Radboud University, Nijmegen, Netherlands
59. TransThera Consulting
60. Duke University, Durham, NC
61. Wolfson Institute of Population Health, Queen Mary University of London, UK
62. Philipps-University Marburg, Germany
63. University of Lagos, Nigeria

