## Supplement Materials for "Neuronal alpha-Synuclein Disease stage progression over five years"

**SUPPLEMENTARY MATERIAL**

Table of Contents

Supplementary Table 1. Staging anchors for application of the NSD-ISS

Supplementary Table 2. Clinical and Biological Baseline Characteristics - Omitted Groups

Supplementary Figure 1. Longitudinal staging of the PPMI NSD cohort (completers only LOCF)

Supplementary Table 3. Tracks leading to stage progression (within 3 years).

**Supplementary Table 1.** **Staging anchors for application of the NSD-ISS**

|  | **Biologic anchors** | | | **Anchors of clinical signs or symptoms (stages 2A and 2B) and functional impairment (stages 3-6)**^1, 2^ | |
| --- | --- | --- | --- | --- | --- |
| **Stage** | **S** | **D** ^a^ | **G** | **Domain** | **Anchor(s)** |
| Stage 0 | - | - | *SNCA* ^b^ | — | — |
| Stage 1A | + | - | ± | (1) Cognitive  (2) Motor  (3) Other non-motor | (1) MDS-UPDRS item 1.1 = 0; and  (2a) Does not have subthreshold parkinsonism ^c^; and (2b) is not on PD medication ^d^; and  (3a) Does not have RBD; and (3b) is not hyposmic ^e^ |
| Stage 1B | + | + | ± |  |  |
| Stage 2A | + | - | ± | (1) Cognitive  (2) Motor  (3) Other non-motor | (1) Item 1.1 = 1 AND MoCA ≥ 25; or  (2a) Has subthreshold parkinsonism ^c^; or (2b) is on PD medication ^d^; or  (3a) Has RBD; or (3b) is hyposmic ^e^ |
| Stage 2B | + | + | ± |  |  |
| Stage 3 | + | + | ± | (1) Cognitive  (2) Motor | (1a) Item 1.1 = 1 AND MoCA ≤ 24; or (1b) Item 1.1 = 2 AND MoCA ≥ 25; or  (2) MDS-UPDRS-II = 3-13 AND either subthreshold parkinsonism ^c^ or PD medication ^d^ |
| Stage 4 | + | + | ± | (1) Cognitive  (2) Motor  (3) Other non-motor | (1a) Item 1.1 = 2 and MoCA ≤ 24; or (1b) item 1.1 = 3 AND MoCA ≥ 25; or  (2) MDS-UPDRS-II = 14-26; or  (3) MDS-UPDRS-I (excluding item 1.1) = 13-24 ^f^ |
| Stage 5 | + | + | ± | (1) Cognitive  (2) Motor  (3) Other non-motor | (1a) Item 1.1 = 3 AND MoCA ≤ 24; or (1b) item 1.1 = 4 AND MoCA ≥ 25; or  (2) MDS-UPDRS-II = 27-39; or  (3) MDS-UPDRS-I (excluding item 1.1) = 25-36 |
| Stage 6 | + | + | ± | (1) Cognitive  (2) Motor  (3) Other non-motor | (1) Item 1.1 = 4 AND MoCA ≤ 24; or  (2) MDS-UPDRS-II ≥ 40; or  (3) MDS-UPDRS-I (excluding item 1.1) ≥ 37 |

**^1^** Presence of qualifying signs/ symptoms in any single domain qualifies for stage 2 but individuals can have combination in all 3 domains.

^2^ Presence of qualifying functional impairment in any single domain qualifies for stage 3-6 but individuals can have combination in all 3 domains.

^a^ D positivity defined as < 75% age/sex-expected lowest putamen SBR.

^b^ Only fully penetrant pathogenic *SNCA* variants qualify for Stage 0.

^c^ Subthreshold parkinsonism defined as MDS-UPDRS-III ≥ 5 excluding postural and action tremor.

^d^ Medication for treating the symptoms of PD as per MDS-UPDRS item 3a

^e^ Hyposmia defined as UPSIT percentile ≤ 15 (age and sex adjusted).

^f^ MDS-UPDRS-I (excluding item 1.1) ≥ 13 is sufficient for stage 4 provided that stage 2 criteria are met.

**Supplementary Table 2. Clinical and Biological Baseline Characteristics - Omitted Groups**

|  | | **NSD Stage at Baseline** | | | |
| --- | --- | --- | --- | --- | --- |
|  | **All Participants** (N = 576) | **Stage 1A** (N = 10) | **Stage 1B** (N = 2) | **Stage 5** (N = 4) | **Stage 6** (N = 1) |
| **Age (years),** Mean (SD) | 61.8 (9.6) | 64.0 (8.7) | 67.3 (NA) | 58.7 (10.6) | 49.6 (NA) |
| **Sex (male),** n (%) | 368 (64%) | 5 (50%) | 2 (NA) | 3 (NA) | 0 (NA) |
| **Years from dx,** Median (Q1, Q3) | 0.5 (0.2, 1.6) | NA | NA | 2.1 (1.8, 5.9) | 4.0 (NA) |
| **MDS-UPDRS Item 1.1 Score,** Median (Q1, Q3) | 0.0 (0.0, 1.0) | 0.0 (0.0, 0.0) | 0.0 (NA) | 3.0 (2.0, 3.0) | 0.0 (NA) |
| **MDS-UPDRS Part I,** Median (Q1, Q3) | 5.0 (3.0, 9.0) | 3.5 (0.0, 6.0) | 3.5 (NA) | 15.0 (11.5, 18.5) | 17.0 (NA) |
| **MDS-UPDRS Part II,** Median (Q1, Q3) | 5.0 (2.0, 8.0) | 0.0 (0.0, 1.0) | 1.0 (NA) | 9.5 (3.5, 22.0) | 40.0 (NA) |
| **MDS-UPDRS Part III (ON),** Median (Q1, Q3) | 17.0 (10.0, 24.0) | 0.5 (0.0, 2.0) | 0.0 (NA) | 11.5 (7.5, 22.5) | 16.0 (NA) |
| Missing | 6 | 0 | 0 | 0 | 0 |
| **MDS-UPDRS Total Score (ON),** Median (Q1, Q3) | 28.0 (19.0, 39.0) | 5.0 (3.0, 7.0) | 4.5 (NA) | 43.0 (24.5, 61.0) | 73.0 (NA) |
| Missing | 6 | 0 | 0 (NA) | 0 | 0 |
| **On PD Medication*,** n (%) | 131 (23%) | 0 | 0 (NA) | 3 (NA) | 1 (NA) |
| **Total LED** Median (Q1, Q3) | 500.0 (300.0, 830.0) | NA | NA | 940.0 (750.0, 1190) | 692.0 (NA) |
| **MOCA Total Score,** Mean (SD) | 26.9 (2.7) | 27.4 (3.5) | 27.5 (NA) | 19.0 (5.9) | NA |
| Missing | 2 | 0 | 0 | 0 | 1 |
| **UPSIT Percentile ≤ 15%,** n (%) | 458 (80%) | 0 | 0 (NA) | 4 (NA) | 1 (NA) |
| Missing | 7 | 0 | 0 | 0 | 0 |
| **Mean Striatum Binding,** Mean (SD) | 1.46 (0.50) | 2.88 (0.38) | 1.56 (NA) | 1.23 (0.23) | 0.84 (NA) |
| **Age/Sex-Expected DAT,** Median (Q1, Q3) | 0.33 (0.25, 0.43) | 1.11 (1.01, 1.23) | 0.57 (NA) | 0.37 (0.26, 0.44) | 0.15 (NA) |
| **Low CSF A-beta 1-42 (<683 pg/mL),** n (%) | 187 (33%) | 3 (30%) | 1 (NA) | 1 (NA) | NA |
| Missing | 10 | 0 | 0 | 0 | 1 |
| **High CSF t-tau (>266 pg/mL),** n (%) | 38 (7%) | 0 | 1 (NA) | 2 (NA) | NA |
| Missing | 6 | 0 | 0 | 0 | 1 |
| **High CSF p-tau (>24 pg/mL),** n (%) | 32 (6%) | 0 | 0 (NA) | 0 (NA) | NA |
| Missing | 6 | 0 | 0 | 0 | 1 |
| **Serum NFL,** Median (Q1, Q3) | 11.7 (8.6, 16.0) | 9.2 (8.9, 10.8) | 16.8 (NA) | 22.3 (11.4, 40.6) | 31.6 (NA) |
| Missing | 53 | 2 | 0 | 1 | 0 |
| **Serum Urate,** Mean (SD) | 313.0 (80.6) | 298.3 (66.1) | 387.0 (NA) | 285.3 (50.2) | 124.0 (NA) |
| Missing | 24 | 2 | 0 | 0 | 0 |
| **No. APOE e4 alleles,** n (%) |  |  |  |  |  |
| 0 | 430 (75%) | 7 (NA) | 1 (NA) | 2 (NA) | 1 (NA) |
| 1 | 132 (23%) | 2 (NA) | 1 (NA) | 2 (NA) | 0 (NA) |
| 2 | 11 (2%) | 0 | 0 (NA) | 0 (NA) | 0 (NA) |
| Missing | 3 | 1 | 0 | 0 | 0 |

*Based on enrollment rules, genetic cohorts were allowed to be on PD medication at baseline.

**Supplementary Figure 1. Longitudinal staging of the PPMI NSD cohort (completers only LOCF)**

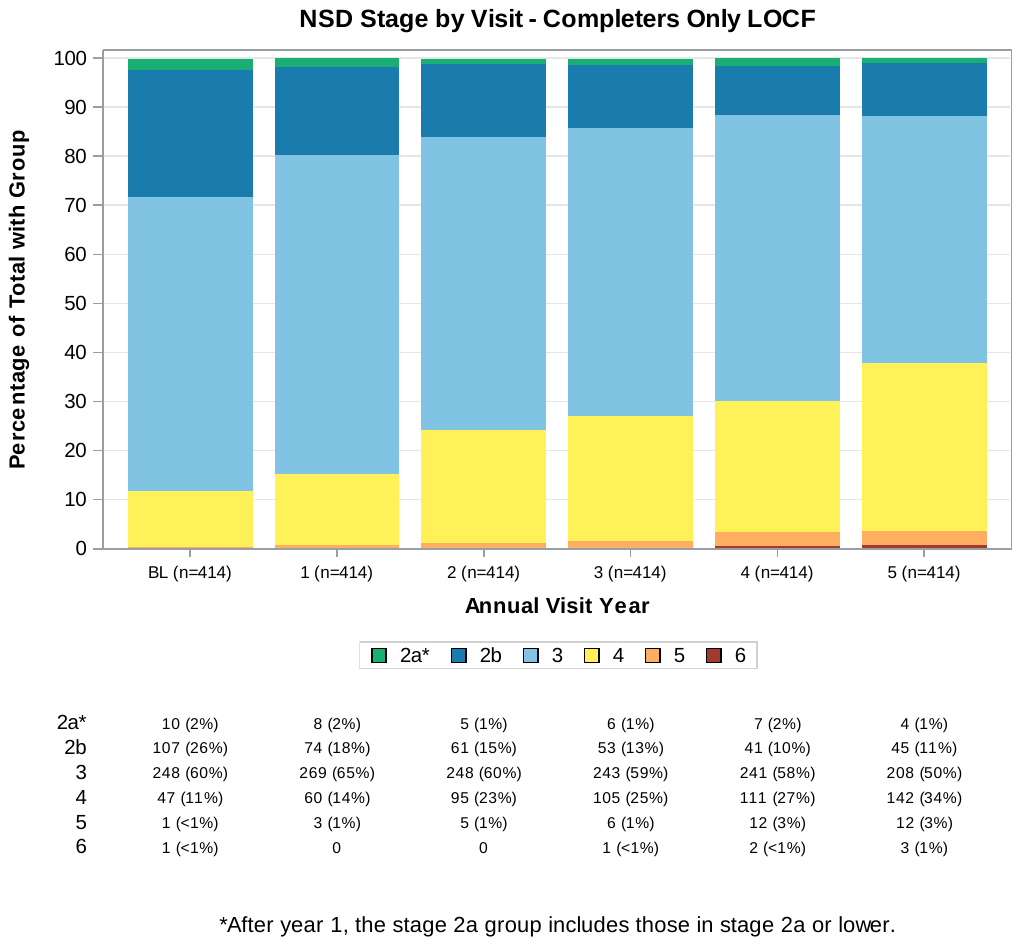

Report Generated on Data Submitted as of: 05Feb2024.

Abbreviations: LOCF= last observation carried forward ; NSD = neuronal a-synuclein disease; NSD-ISS = neuronal a-synuclein disease integrated staging system.

* After Year 1, Stage 2A group includes those in Stage 2A or lower.

** Excludes participants who are Stage 0, 1A and 1B (N=13) at baseline; participants who are Stage 2A at Baseline and have no follow up DAT (N=8)

**Supplementary Table 3. Tracks leading to stage progression (within 3 years).**

|  | **Subgroup** | | |
| --- | --- | --- | --- |
| **Track** | **Progressed to Stage 3 ^a^** (N=92) | **Progressed to Stage 4 ^b^** (N=137) | **Progressed to Stage 5 ^c^** (N=11) |
| **Years to first progression**, mean (SD) | 1.5 (0.8) | 1.9 (0.8) | 1.7 (0.8) |
| Median (IQR) | 1.1 (1.0, 2.0) | 2.0 (1.0, 3.0) | 2.0 (1.0, 2.0) |
| **Met criteria for track** |  |  |  |
| Cognitive | 9 (10%) | 28 (20%) | 6 (55%) |
| Motor | 87 (95%) | 75 (55%) | 4 (36%) |
| Non-Motor | N/A | 69 (50%) | 2 (18%) |
| **Combination of tracks** |  |  |  |
| Cognitive only | 5 (5%) | 18 (13%) | 6 (55%) |
| Motor only | 83 (90%) | 47 (34%) | 3 (27%) |
| Other Non-Motor only | N/A | 42 (31%) | 1 (9%) |
| Cognitive + Motor | 4 (4%) | 3 (2%) | N/A |
| Motor + Other Non-Motor | N/A | 20 (15%) | 1 (9%) |
| Cognitive + Other Non-Motor | N/A | 2 (1%) | N/A |
| Cognitive + Motor + Other Non-Motor | N/A | 5 (4%) | N/A |

a Reflects participants whose first progression was to stage 3. In most cases, these are individuals who started in stage 2B and progressed to stage 3.

b Reflects participants whose first progression was to stage 4. In most cases, these are individuals who started in stage 3 and progressed to stage 4.

c Reflects participants whose first progression was to stage 5. In most cases, these are individuals who started in stage 4 and progressed to stage 5.
